# Effects of water quality, sanitation, handwashing and nutritional interventions on enteric pathogen seroconversion in rural Bangladesh: a cluster randomised trial

**DOI:** 10.64898/2026.10.30.26366885

**Authors:** Mahbubur Rahman, Pearl Anne Ante-Testard, Richelle W. Kihoro, Henry Kanyi, Henry Emisiko, Huiyu Hu, Everlyn Kamau, E. Brook Goodhew, Chabier Coleman, Shahjahan Ali, Md Ziaur Rahman, Abul Kasham Shoab, Md. Rana Miah, Sania Ashraf, Sarker M Parvez, Md. Mostofa Shakil, Samantha J. Bents, Julianna Colado, Nikolina Walas, Jessica A. Grembi, Richelle C. Charles, Edward T. Ryan, Claire Munroe, Robert L. Atmar, Jade Benjamin-Chung, Ayse Ercumen, Andrew N. Mertens, Audrie Lin, Patrick J. Lammie, Christine P. Stewart, John M. Colford, Stephen P. Luby, Sammy M. Njenga, Rashidul Haque, Jeffrey W. Priest, Benjamin F. Arnold

## Abstract

**Background:** Improvements to water quality, sanitation, handwashing (WASH) and nutrition may prevent enteric pathogen infection during the first two years of life. Few randomised controlled trials have measured pathogen-specific effects of WASH and nutritional interventions.

**Methods:** Within a 2×2 factorial cluster-randomized trial of WASH and nutritional interventions in Bangladesh, we tested dried blood spot specimens with multiplex assay for IgG responses to 12 enteric pathogens: *Norovirus*, *Campylobacter*, LT-producing enterotoxigenic *E. coli* (LT-ETEC), *Shigella*, non-typhoidal and typhoidal *Salmonella enterica*, *Vibrio cholerae*, *Cryptosporidium*, *Giardia*, *Entamoeba histolytica*, *Taenia solium*, and *Strongyloides stercoralis*. Primary analysis included children with IgG measurements at ∼24 months, and secondary analysis assessed early effects in a sub-study at ∼14 months. We estimated age-specific seroprevalence and marginal seroprevalence differences for WASH and nutrition effects. Trial is registered with ClinicalTrials.gov (NCT01590095).

**Findings:** Across the four arms (450 clusters), 4817 samples were tested for IgG responses, 2453 samples at the primary endpoint. Seroprevalence increased with age, consistent with intense early-life transmission. For highest transmission pathogens, >94% of children had seroconverted to *norovirus*, LT-ETEC and *Campylobacter* by 12-18 months. WASH effects were modest and pathogen-specific but generally protective. At 24 months, children that received WASH interventions had less antibody against *Shigella* (–4.5%, 95% CI –7.6% to –1.5%), non-typhoidal *S. enterica* (–4.0%, 95% CI –6.5% to –1.5%), and *Giardia* (–3.4%, 95% CI –6.7% to –0.2%), although not after multiple-testing correction. Nutrition had limited effects on most enteric pathogens, but reduced non-typhoidal *S. enterica* at 14 months (–6.4%, 95% CI: –9.8% to –3.0%).

**Interpretation:** Multiplex serology provided evidence that WASH may modestly reduce cumulative exposure to selected enteric pathogens, but household-level interventions were insufficient to substantially disrupt transmission in this high-exposure setting.

**Funding:** National Institutes of Health (R01AI166671, R01AI106878, R01AI134814). The original WASH Benefits trial was funded by Gates Foundation (OPPGD759).

## Introduction

Enteric pathogens are a leading cause of the global burden of disease, with the greatest burden falling on young children in low-resource settings.^1^ Although this burden is often recognised through diarrhoeal illness and diarrhoea-associated mortality, many enteric infections in early childhood are subclinical.^2^ These infections may nevertheless have lasting consequences, including malnutrition, linear growth faltering,^3^, and impaired cognitive development.^5^ Preventing exposure is difficult because enteric pathogens move through overlapping transmission pathways, including waterborne, foodborne, person-to-person, and other faecal-oral routes that vary across social and ecological contexts. Nutrition may also influence infection risk, as undernutrition can compromise gut barrier function and increase vulnerability to enteric infection.^6^ As a result, it remains difficult to determine which pathogens may be most affected by water, sanitation, and handwashing (WASH), and nutrition interventions, and at what point in early childhood those effects emerge.

The WASH Benefits Bangladesh cluster-randomised trial provided an important opportunity to examine these questions. The trial evaluated individual and combined WASH and nutrition interventions on child growth and diarrhoea, with children followed from birth to approximately 24 months.^7,8^ Prior analyses showed that combined WASH interventions reduced caregiver-reported diarrhoea by 31-38%, *Giardia* infections by 27-30% (aged ∼31 months),^9^ and soil transmitted helminth infection by 10% (∼30 months for index children and 7 years for non-index children)^10^ using stool-based testing. Quantitative polymerase chain reaction (PCR) testing in a sub-study at age ∼14 months demonstrated that the WASH interventions reduced viral pathogen infections, but found no differences in bacterial or protozoan pathogens.^11^ These results suggest that WASH may interrupt multiple faecal-oral transmission pathways, but stool-based testing at limited time points provides an incomplete picture of cumulative pathogen exposure and intervention effects over time. A richer characterization of pathogen-specific transmission could help clarify whether WASH affected enteric infection in Bangladesh and why this trial showed stronger intervention effects than contemporaneous trials of similar interventions fielded in Kenya and Zimbabwe.^12,13^

Here, we report results of multiplex IgG antibody testing for 12 enteric pathogens in the Bangladesh birth cohort. Our objective was to estimate the effects of improved WASH and nutrition interventions on enteric pathogen seroconversion during the first two years of life, a key window for enteric pathogen seroconversion in high transmission settings.^14^ Because IgG responses integrate exposure over time, serology offers a complementary approach for measuring pathogen-specific force of infection in trials with infrequent specimen collection.^15^ We hypothesized that WASH could reduce seroconversion by interrupting environmental and person-to-person transmission. We also hypothesized that the nutrition intervention could reduce seroconversion through two mechanisms: exclusive breastfeeding through six months followed by continued breastfeeding with complementary food through 24 months could reduce enteric pathogen exposure and, if exposed, may lead to a more robust innate immune response, pre-empting a systemic IgG response. We hypothesized that there would be no interaction between WASH and nutrition interventions for most of the pathogens. A companion trial conducted in Kenya addressed the same study objectives.[Kanyi et al., 2026, preprint]

## Methods

### Study design and participants

The WASH Benefits Bangladesh study was a seven arm, cluster-randomised trial conducted in rural districts of Gazipur, Mymensingh, Kishoreganj, and Tangail in central Bangladesh (**figure 1a**). The study targeted communities with low groundwater iron and arsenic and no major ongoing or planned WASH or nutrition programs. The area is predominantly agricultural, and households are commonly organised into compounds where multiple families share a courtyard and may also share a pond, water source, and latrine. Pregnant women in their second or third trimester were eligible to enrol if they planned to remain in the study village for the following two years, regardless of where they delivered. Only one pregnant woman was enrolled per compound; in the case of twin births, both children were enrolled. Details of the study design and rationale^7^, and primary outcomes results on diarrhoea and child growth^8^ have been published previously. Study enrolment occurred between May 2012 and July 2013.

**Figure 1.**
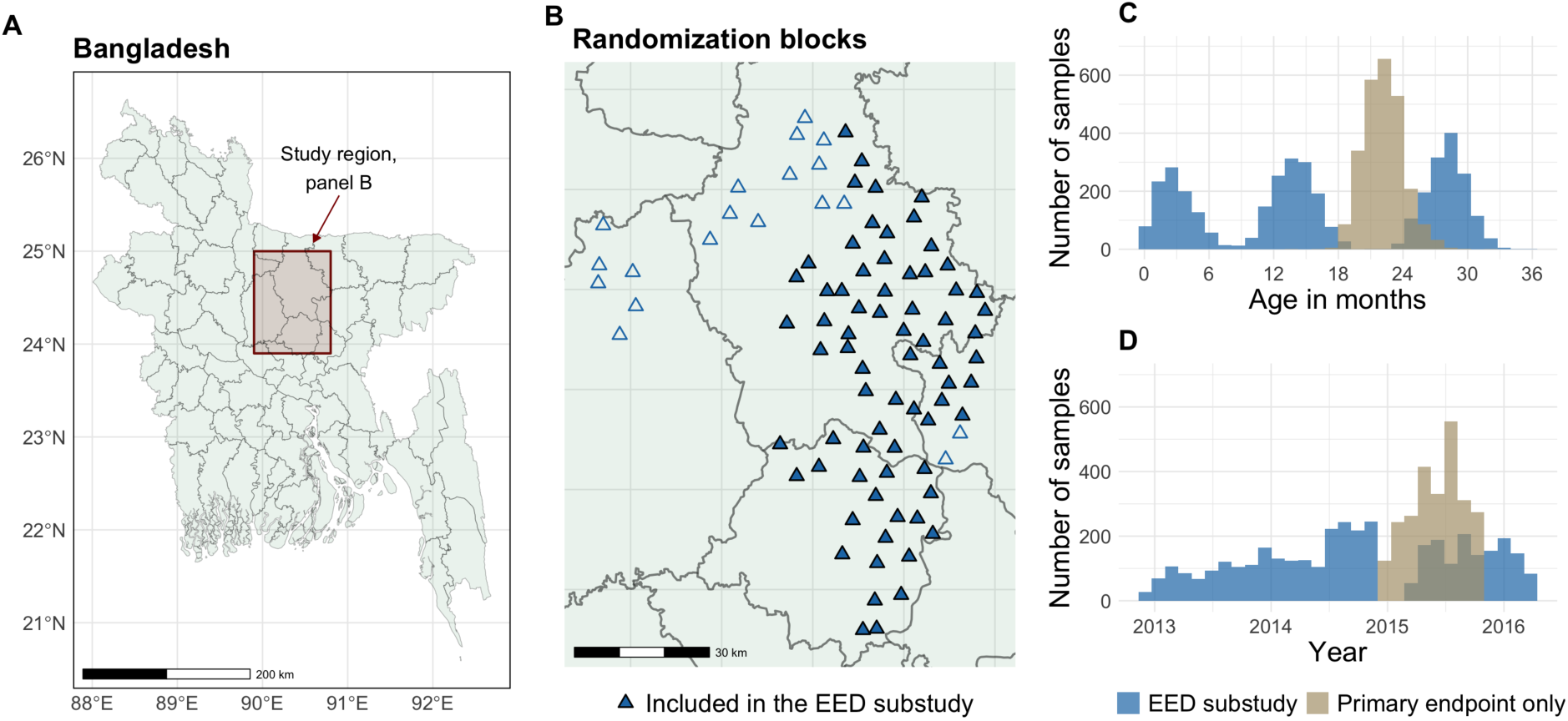
Overview of the WASH Benefits Bangladesh trial serology study. **A**. Trial location within Bangladesh. **B.** Location of 90 randomisation blocks in the trial. Each block included 8 village clusters, with five allocated to a double-sized Control, WASH, Nutrition, and WASH+Nutrition. Of 90 randomization blocks, 71 were included in the Environmental Enteric Dysfunction (EED) substudy with children measured longitudinally at up to three visits. **C.** Distribution of blood samples tested in the control, WASH, nutrition, and WASH+Nutrition arms by age for 1464 children in the EED substudy at three visits (median ages at 3, 14, 28 months) and remaining 989 children measured once during their final visit (median age at 22 months). EED and non-EED children were included in the primary analysis, total of 2453 samples (median age at 24 months) **D.** Distribution of samples in panel C by calendar date rather than child age.

The trial included six intervention arms: water treatment (W), sanitation upgrades (S), handwashing promotion (H), nutrition, combined WASH, combined WASH + Nutrition, and a double-sized control arm. The water intervention included chlorine tablets and a safe drinking water storage vessel. The sanitation intervention included improvements via a double-pit latrine with a water seal, child potties, and hoes for faeces disposal at the compound level. The handwashing intervention included handwashing stations placed near the latrine and kitchen, stocked with detergent soap and dispensers. Within the combined intervention arms, the sanitation intervention was implemented first, followed sequentially by handwashing and water treatment. The nutrition intervention consisted of age-appropriate infant and young child feeding counselling plus lipid-based nutrient supplements provided twice daily from 6 to 24 months of age. Interventions were initiated during pregnancy so that children were exposed from birth.

The trial’s environmental enteric dysfunction (EED) sub-study assessed the effects of interventions on EED among a subsample of children who underwent intensive biological sampling^16^. EED sub-study clusters were selected according to the feasibility of specimen collection and transport to a central laboratory. The EED sub-study included children in the control, combined WASH, Nutrition, and combined WASH + Nutrition. For this analysis, we used a 2×2 factorial design across these four arms to preserve statistical power and maintain a consistent definition of the WASH intervention package.

### Randomisation and masking

In the WASH Benefits Bangladesh trial, compounds were enrolled within 720 clusters, grouped into geographically matched blocks of eight clusters^7^ (**figure 1a and 1b**). Within each block, the trial randomly allocated clusters to one of the seven arms. For the EED sub-study, a total of 450 clusters were randomly allocated to the four arms: WASH, WASH + Nutrition, Nutrition and double-sized control arm.

Because the interventions had distinct visible components, neither participants nor field staff could be masked to assignment; however, data collection and intervention delivery were conducted by separate teams. Here, to minimise analytic bias, an investigator (PAA-T) performed masked statistical analyses from the raw datasets, using a re-randomised, uninformative assignment variable in place of the true group labels until final estimates were generated.

### Ethics

The WASH Benefits trial (ClinicalTrials.gov identifier: NCT01590095) received ethical approval from multiple institutional review boards, including the Ethical Review Committee of the International Centre for Diarrhoeal Disease Research, Bangladesh (PR11063); the Committee for the Protection of Human Subjects at the University of California, Berkeley (2011-09-3652); the Institutional Review Board at Stanford University (25863); and the Institutional Review Board at the University of California, San Francisco (22-36722).

Procedures

#### Sample collection and testing

Children in the EED sub-study provided longitudinal blood specimens at three visits, with median ages of approximately 3, 14, and 28 months^16^, from December 2012 after intervention initiation through March 2016 (**figure 1**). In addition, dried blood spots (DBS) were collected from non-EED children in the birth cohort at the final study visit, at a median age of approximately 22 months. For the primary endpoint analysis, we included all children with available DBS at the final study visit, combining EED children measured at visit 3 with non-EED children measured once at their final visit; the overall median age at this endpoint was approximately 24 months. Throughout the paper, we refer to visit 3 or final visit analysis as the 24-month/primary analysis and visit 2 as the14-month/secondary analysis.

DBS were collected on TropBio filter paper, with six extensions, each calibrated to absorb approximately 10 μL of whole blood. These were then dried at ambient temperature overnight, individually sealed with desiccant in plastic bags, and stored at −80°C at the International Centre for Diarrhoeal Research, Bangladesh (icddr,b) before being shipped to the Kenya Medical Research Institute (KEMRI) at ambient temperature and stored at −80°C.

One dried blood spot extension of approximately 10 μL from each participant was eluted to 1:40 overnight at 4°C with 200 µl of Buffer B (containing 1X phosphate buffer saline (PBS), 0.50% polyvinyl alcohol (PVA), 0.80% polyvinylpyrrolidone (PVP), 0.30% Tween-20, 0.02% sodium azide, 0.50% casein and 3 µg/mL *Escherichia coli* extract). Following elution, samples were diluted in Buffer B to a final serum dilution of 1:400 prior to analysis using a multiplex bead-based assay.^17^

Serum dilutions were centrifuged in a microcentrifuge at 4,000 RPM for 5 minutes immediately before use to pellet any residual *E. coli* extract particulates. Bloodspot dilutions were then incubated with antigen-coupled microsphere beads, and fluorescence was measured using a Bio-Plex 200 instrument equipped with Bio-Plex Manager 6.1 software (Bio-Rad, Hercules, CA). Targets (antigen-coupled beads) included *Norovirus* (GI.5 and GII.3, GII.4, GII.6, GII.17 virus-like particles), *Campylobacter* (p18, p39), LT-producing enterotoxigenic *E. coli* (LT-ETEC), *Shigella*/enteroinvasive *E. coli* (EIEC) (IpaB), *Salmonella enterica* serovar *Typhimurium* (LPS Group B), and *S. enterica* serotypes Enteritidis and *S. enterica* serovars Typhi/Paratyphi A (HlyE) (LPS Group D), *Vibrio cholerae* (OSP-Inaba, OSP-Ogawa), *Cryptosporidium* (Cp17, Cp23), *Giardia* (VSP-5), *Entamoeba histolytica* (LecA), *Taenia solium* (Es33, T24), and *Strongyloides stercoralis* (NIE). *Schistosoma japonicum* (GST) was included as a negative control protein. A detailed description of the laboratory procedures has been published previously.^18^

Each assay plate contained duplicate wells for Buffer B-only to serve as blanks, as well as one negative control serum dilution and four pooled positive control sera with known antibody reactivity at varying concentrations for quality control purposes. Results were expressed as median fluorescence intensity minus background (MFI-bg).

Quality assurance procedures included monitoring positive control performance using Levey-Jennings plots across 77 plates; plates were repeated if control values fell outside ±2 standard deviations of the established mean. Control samples were run in duplicate to evaluate within-plate and between-plate variability. Plates were flagged for repeat testing if control coefficients of variation exceeded 20% or if bead counts were consistently low (fewer than 20 beads per analyte).

#### Seropositivity cutoffs

Antigen-specific seropositivity cutoffs were defined using Receiver-Operating-Characteristic (ROC) analysis of known positive/negative reference specimens for protozoan and helminth targets (Cp17, Cp23, VSP-5, LecA, Es33, T24) or using a longitudinal pre-exposed cohort approach for bacterial and viral pathogens.^19^ In the pre-exposed cohort approach, we used IgG responses from all timepoints measured and identified incident IgG increases of ≥100-fold from an initial low value (≤300 MFI-bg). We enforced the large increase between measurements to reduce the possibility of including measurements that were mid-boost since the timing of blood collection was independent of infection. Pre-boost values were taken to represent the seronegative distribution, and cutoffs were set as mean + 3 SD of pre-boost log10 IgG (imputing MFI-bg ≤0 to 1). Cutoffs were derived using both WASH Benefit trial cohorts (Bangladesh, Kenya) because samples were processed identically in the same lab using the same bead couplings and reagents (**supplementary figure 1)**. For pathogens with multiple antigens in the assay, children were classified as seropositive if positive for any antigen.

### Outcomes

The primary outcome was IgG seroprevalence at visit 3 of all eligible children (∼median age 24 months) (**figures 1c and 1d**). We evaluated seroprevalence for the 12 enteric pathogens: *Norovirus* GI/GII, *Campylobacter*, LT-ETEC, *Shigella*/EIEC, non-typhoidal *S. enterica*, typhoidal *S. enterica*, *V. cholerae*, *Cryptosporidium*, *Giardia*, *E. histolytica*, *T. solium*, and *S. stercoralis*. Seroprevalence at visits 2 and 3 was interpreted as a measure of cumulative incidence of seroconversion. Enteric pathogen seroprevalence and seroconversion rates coincide in this narrow age range during the window primary infection in high transmission settings since IgG responses reflect past exposure and persist beyond acute infection.^14,20^ Secondary outcomes included IgG seroprevalence at visit 2 (∼median age 14 months) among children enrolled in the longitudinal EED sub-study, and the count of seropositive pathogens per child (range of 0 to 12).

### Statistical analyses

Sample size was determined by the number of available banked specimens. For pooled factorial comparisons, we estimated minimum detectable effects assuming 80% power, two-sided significance level of 0.05, an intracluster correlation coefficient of 0.15, and 110 clusters per comparison group, with an average of eight children per cluster. Conservatively assuming 50% seroprevalence in the control group, the minimum detectable relative reduction was approximately 20%, corresponding to an absolute reduction of 10 percentage points. These calculations were based on the number of clusters in the EED sub-study and did not incorporate potential gains in precision from geographic matching.^21^

We conducted an intention-to-treat analysis based on the 2×2 factorial design: WASH (WASH and WASH + Nutrition) versus no WASH (Nutrition and control) and Nutrition (Nutrition and WASH + Nutrition) versus no Nutrition (WASH and control). Across all samples tested, we estimated age- and visit-specific IgG seroprevalence by intervention group. We conducted a complete-case analysis in which the primary analysis was restricted to children with available DBS specimens at the final study visit, at a median age of 24 months, and the secondary analysis was restricted to EED sub-study children with available DBS specimens at visit 2, at a median age of 14 months.

We estimated marginal average seroprevalence and risk differences at visit 3 (median age 24 months) with a linear regression model with main effects for WASH and Nutrition interventions and indicators for randomization blocks. We used clustered-robust standard errors to account for repeated measurements within each study cluster. We assessed WASH x Nutrition interaction with an interaction term in the model and if there was no evidence of additive interaction (P>0.1, pre-specified), we pooled arms to estimate each main effect separately to improve precision. The main effect for each intervention was averaged over the levels of the other intervention. Conditional means and effects were reported in cases of interaction between WASH and nutrition interventions. We computed two-sided permutation p-values by permuting assignments within randomization blocks (10,006 replicates) and controlled the false discovery rate across 12 pathogens using a Benjamini-Hochberg approach allowing for 5% false discovery.

We repeated the analysis at visit 2 among EED sub-study children (median age 14 months) to assess early intervention effects. The EED sub-study was not balanced by randomisation block, so models excluded block indicators but otherwise followed the primary analysis approach. Lastly, we estimated differences between groups in the number of pathogens seropositive at the final visit using linear regression with WASH and Nutrition indicators, randomization-block indicator, and cluster-robust standard errors, testing for interaction and pooling arms when appropriate as in the primary analysis.

## Results

### Study population and characteristics

The trial randomised 720 clusters and enrolled 5551 pregnant women living in 5551 compounds. The pre-specified analysis of IgG outcomes (Zenodo: https://doi.org/10.5281/zenodo.23671136) focused on the four factorial WASH and Nutrition arms, including 450 clusters and a total of 4817 IgG measurements, including 2453 collected at the primary endpoint (median age of 24 months) (**figure 2**). At the primary endpoint, DBS samples were available from 180 clusters (897 children) in the control arm, 90 clusters (534 children) in the WASH arm, 90 clusters (495 children) in the Nutrition arm, and 90 clusters (527 children) in the WASH + Nutrition arm. At the visit 2 secondary endpoint, 1370 IgG measurements were available from children in the EED sub-study. Baseline characteristics were well balanced across intervention groups with minimal differences (**table 1, supplementary table 1**).

**Figure 2:**
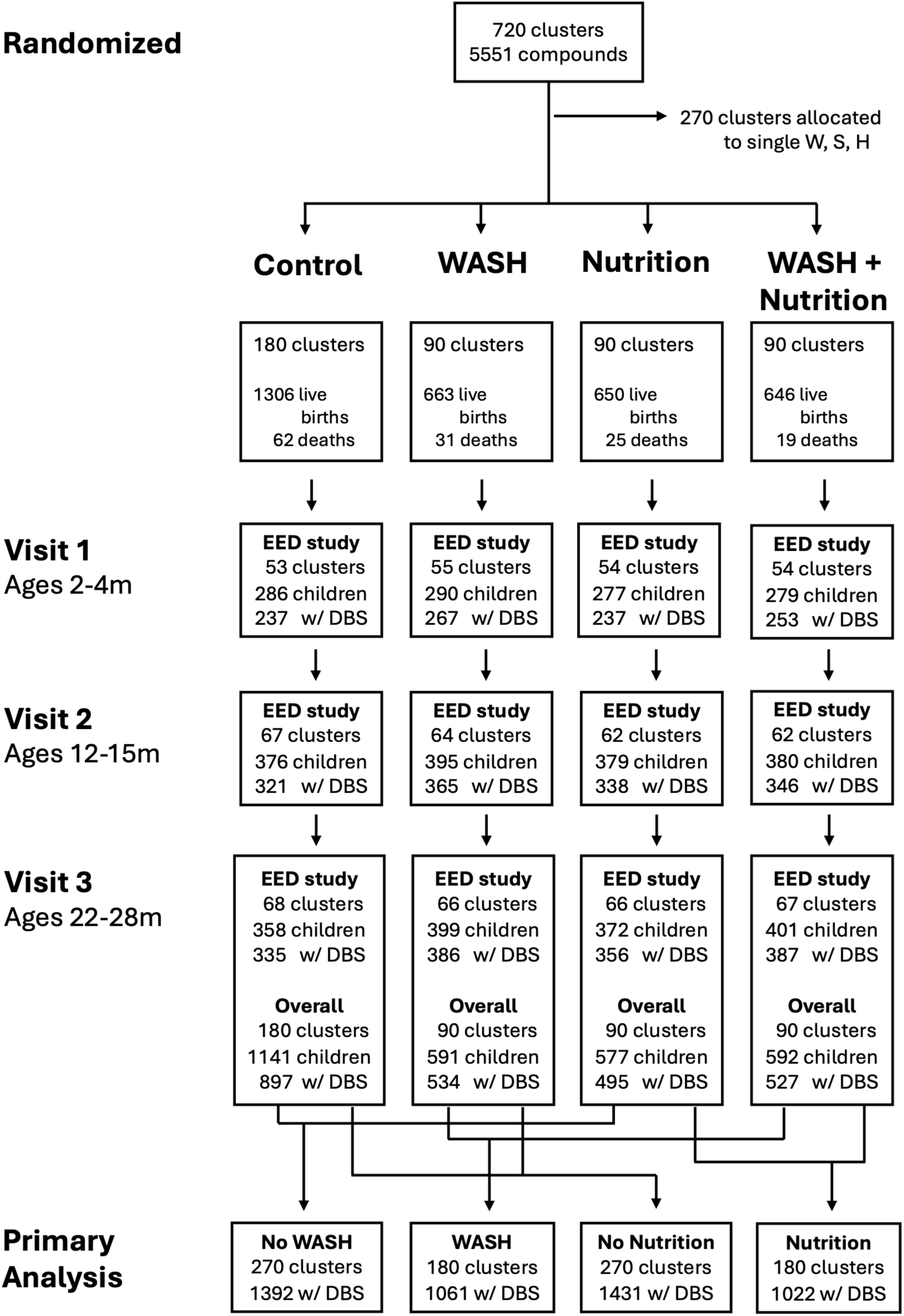
CONSORT flow diagram of clusters and children enrolled in the WASH Benefits Bangladesh enteric pathogen serology study. Ages of measurement indicate the interquartile range at each visit. Overall includes samples from the EED sub-study and non-EED main study. WASH=combined water, sanitation and handwashing; EED=Environmental Enteric Dysfunction; DBS=dried blood spots.

**Table 1:** Baseline characteristics by WASH and Nutrition intervention margins in the factorial design of the WASH Benefits Bangladesh trial. Means by intervention arm are provided in Supplementary Table 1.

|  | **WASH<br>(N=1061)** | **No WASH<br>(N=1392)** | **Nutrition<br>(N=1022)** | **No Nutrition<br>(N=1431)** |
|---|---|---|---|---|
| **Maternal age (years)**<br>Median [Min, Max] | 24 [14, 44] | 23 [15, 45] | 23 [14, 45] | 24 [15, 44] |
| **Maternal education**<br>No education | 152 (14.3%) | 200 (14.5%) | 163 (16.0%) | 189 (13.3%) |
| Primary (1-5y) | 339 (32.0%) | 425 (30.7%) | 312 (30.6%) | 452 (31.7%) |
| Secondary (>5y) | 570 (53.7%) | 759 (54.8%) | 544 (53.4%) | 785 (55.0%) |
| **Maternal height (cm)**<br>Median [Min, Max] | 150 [133, 167] | 151 [127, 168] | 150 [127, 168] | 151 [134, 167] |
| **Paternal education**<br>No education | 321 (30.4%) | 402 (29.1%) | 307 (30.2%) | 416 (29.3%) |
| Primary (1-5y) | 322 (30.5%) | 420 (30.4%) | 325 (32.0%) | 417 (29.3%) |
| Secondary (>5y) | 414 (39.2%) | 559 (40.5%) | 385 (37.9%) | 588 (41.4%) |
| **Works in agriculture** | 322 (30.4%) | 440 (31.8%) | 329 (32.3%) | 433 (30.4%) |
| **International wealth index**<br>Median [Min, Max] | 31.5 [4.66, 63.0] | 31.8 [0, 63.0] | 31.0 [0, 63.0] | 33.7 [0.00, 63.0] |
| **Number of households per compound** Mean (SD) | 11.0 (6.19) | 11.2 (6.30) | 11.3 (6.50) | 11.0 (6.07) |
| **Number of people per compound** Mean (SD) | 4.69 (2.09) | 4.70 (2.18) | 4.71 (2.11) | 4.69 (2.17) |
| **Primary drinking water source (tubewell)** | 815 (76.8%) | 1058 (76.4%) | 758 (74.4%) | 1115 (78.2%) |
| **Distance to primary water source (min)**<br>Mean (SD) | 0.844 (4.06) | 1.05 (16.2) | 0.662 (1.69) | 1.18 (16.2) |
| **Reported storing drinking water** | 475 (44.8%) | 613 (44.3%) | 462 (45.3%) | 626 (43.9%) |
| **Reported treating water today or yesterday** | 2 (0.2%) | 3 (0.2%) | 2 (0.2%) | 3 (0.2%) |
| **Latrine**<br>No latrine | 490 (48.9%) | 627 (48.1%) | 464 (48.4%) | 653 (48.4%) |
| Latrine without water seal | 358 (35.7%) | 428 (32.8%) | 335 (34.9%) | 451 (33.5%) |
| Latrine with water seal | 155 (15.5%) | 249 (19.1%) | 160 (16.7%) | 244 (18.1%) |
| **Primary handwashing location has water and soap** | 240 (24.4%) | 289 (22.6%) | 220 (23.3%) | 309 (23.4%) |
| **HFIAS food insecurity**<br>Moderate/severe | 223 (21.0%) | 305 (22.0%) | 209 (20.5%) | 319 (22.4%) |

WASH=combined water, sanitation and handwashing; SD=standard deviation; HFIAS=Household Food Insecurity Access Scale.

### Intervention adherence

Adherence to the WASH intervention was high and consistent through the entire follow-up period, with markedly higher coverage of WASH indicators in compounds that received WASH interventions (**supplementary table 2**). Adherence to the nutrition intervention was also high, with children in the arms that included the nutrition intervention consuming 93-94% of expected sachets at years 1 and 2 follow-up (**supplementary table 2**).

### Age-specific IgG seroprevalence

IgG seroprevalence curves show that enteric pathogen transmission was intense during the first years of life (**figure 3**). Maternal IgG contributions were evident for many pathogens resulting in a decline in seroprevalence during the first 6-12 months before a subsequent increase due to incident infections. Due to the longevity of IgG responses, seroprevalence curves likely approximate cumulative incidence of seroconversion during this early age window. For the highest transmission pathogens, *Norovirus* GI/GII, LT-ETEC, and *Campylobacter*, over 90% of children had seroconverted by age 24 months and there was no evidence for reductions due to WASH at any age. In contrast, age-seroprevalence curves rose more slowly for several enteric pathogens among children that received improved WASH, reflecting lower transmission due to the WASH intervention. This was most clear for *Shigella/EIEC*, *Giardia* and non-typhoidal *S. enterica* (**figure 3**). Patterns in age-seroprevalence were broadly similar when stratified by the Nutrition intervention (**supplementary figure 3**).

**Figure 3:**
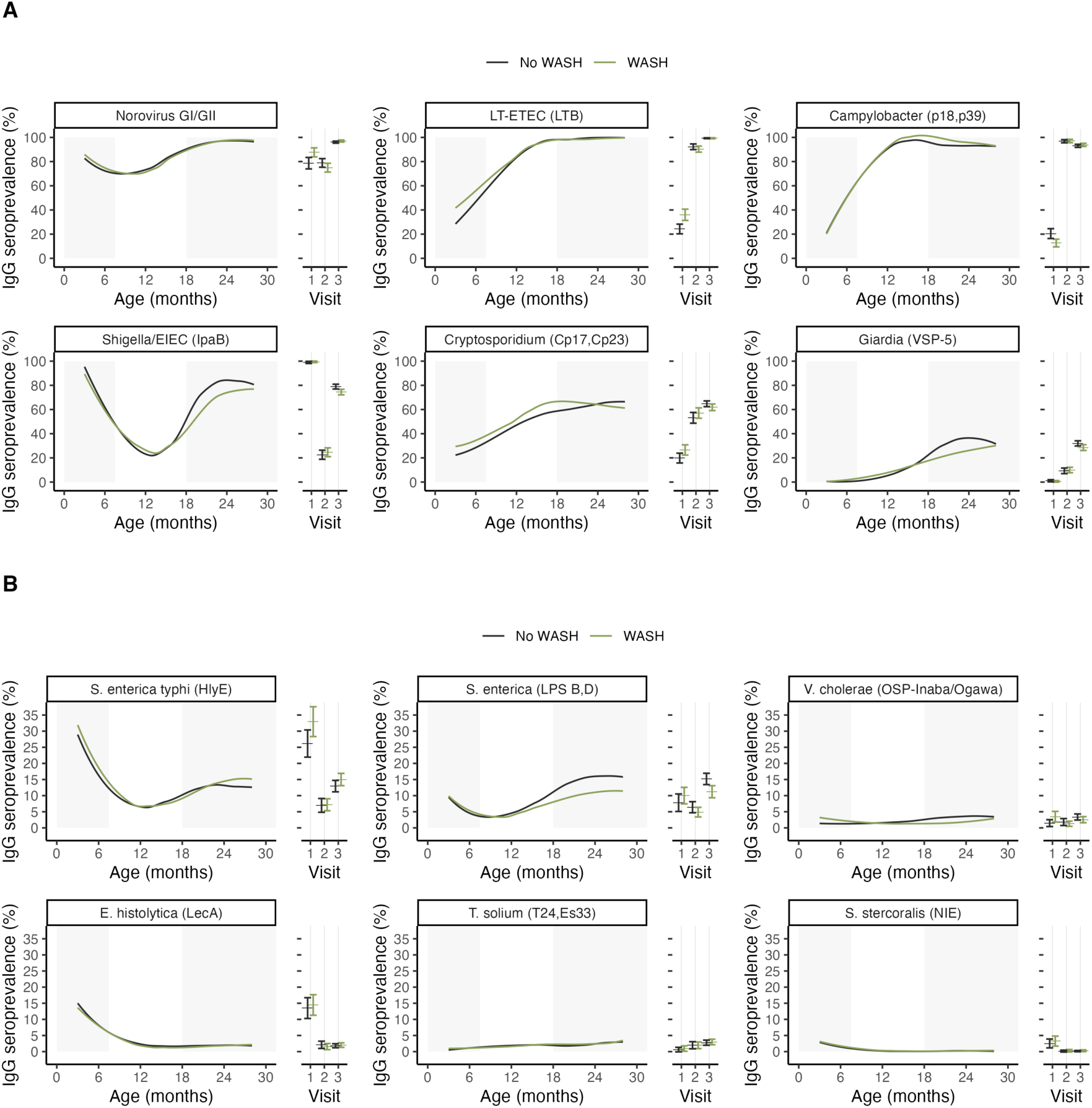
Enteric pathogen seroprevalence by age and WASH intervention group, Bangladesh 2013-2016. **A.** Higher transmission pathogens. **B.** lower transmission pathogens. The difference between panels is the Y-axis range, to better display the data. Lines represent age-seroprevalence estimated by treatment group using locally weighted regression over individual seropositivity data (points), trimmed to ages 3-28 months to avoid edge effects. Points and error bars to the right of each panel show treatment-specific marginal average seroprevalence and 95% confidence intervals by study visit (1, 2, 3). Gray shading shows child age ranges of visits 1 and 3, with the primary outcome and full cohort measured at visit 3. Similar estimates summarized by the Nutrition intervention margin are in **Supplemental Figure 3**. WASH=combined water, sanitation and handwashing intervention.

### Intervention effects at the primary endpoint

For pathogens without evidence of WASH x nutrition interaction, marginal comparisons at the final visit (median age ∼24 months) were consistent with reductions in seroprevalence to *Shigella/*EIEC (– 4.5%, 95% CI –7.6% to –1.5%), non-typhoidal *S. enterica* (–4.0%, 95% CI –6.5% to –1.5%), *V. cholerae* (–0.8%, 95% CI –2.2% to 0.5%), *Cryptosporidium* (–2.9%, 95% CI –6.5% to 0.7%), and *Giardia* (–3.4%, 95% CI –6.7% to –0.2%), though differences were not significant at the 5% level after correcting for multiple testing (**figure 4**). Analyses to estimate the effect of nutrition intervention showed no marked differences between groups in pathogen-specific seroprevalence at the final visit (**supplementary table 3, supplementary figure 2**).

**Figure 4:**
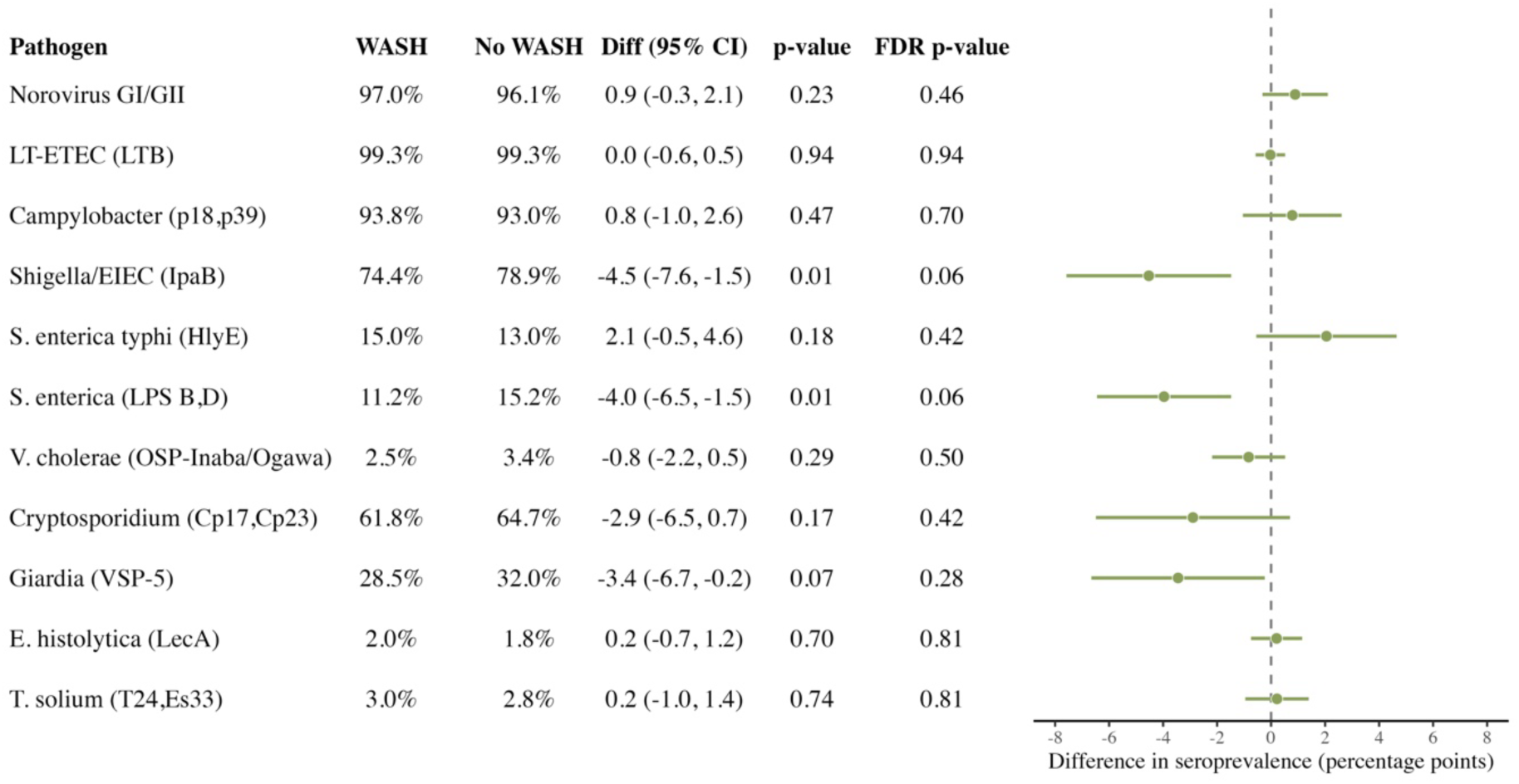
WASH intervention effects on enteric pathogen seroprevalence at median age 24 months, Bangladesh. Effects estimated as marginal means and treatment effects in the factorial design for clusters that received a combined WASH package (WASH, WASH + Nutrition) compared with clusters that did not (Nutrition, Control). Two-sided permutation p-values were estimated by permuting treatment assignments within matched blocks (10,006 replicates). P-values were adjusted using the Benjamini-Hochberg procedure across the 12 comparisons (5% false discovery rate). Excluded results for *S. stercoralis* (NIE) because seroprevalence was low (<1%). WASH=combined water, sanitation and handwashing; Diff=difference; 95% CI=95% confidence interval; FDR=False Discovery Rate.

### Secondary outcomes

Secondary analyses that compared groups at visit 2 (median age 14 months) showed lower seroprevalence to non-typhoidal *S. enterica* among children that received WASH (–5.0%, 95% CI – 8.4% to –1.6%, multiple comparison adjusted p-value = 0.05) but no other marked differences **(figure 5**). For the effect of nutrition, age-seroprevalence curves were consistent with a modest delay in the increase in IgG seroprevalence for *Shigella/*EIEC, *Giardia* and non-typhoidal *S. enterica* (**supplementary figure 3**), with largest conditional effect difference between groups (nutrition vs control) for non-typhoidal *S. enterica* (–6.4%, 95% CI –9.8% to –3.0%, **supplementary table 4, supplementary figure 4**). The number of seropositive enteric pathogens detected at 24 months were broadly similar between children in all intervention groups (median of 5 pathogens seropositive), with slight reductions among children who received the WASH intervention (mean difference –0.1, 95% CI –0.19 to –0.03) (**supplementary figure 5**).

**Figure 5:**
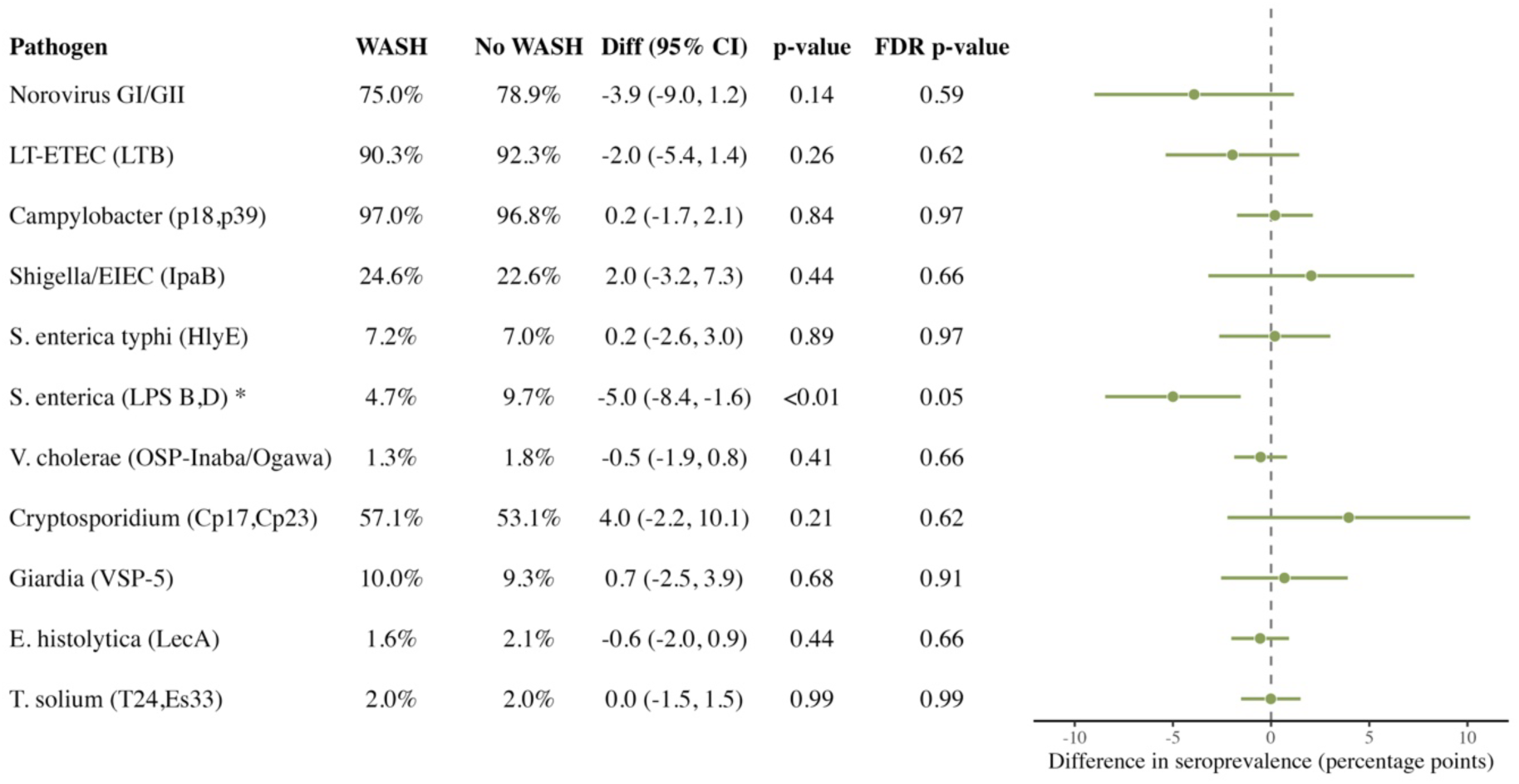
WASH intervention effects on enteric pathogen seroprevalence at median age 14 months, Bangladesh. Effects estimated as marginal means and treatment effects in the factorial design for clusters that received a combined WASH package (WASH, WASH + Nutrition) compared with clusters that did not (Nutrition, Control). *Conditional means and effects comparing WASH vs Control are reported in cases of interaction between WASH and Nutrition interventions (P< 0.1) for *S. enterica* LPS Groups B and D. Two-sided permutation p-values were estimated by permuting treatment assignments (10,006 replicates). P-values were adjusted for multiple comparisons using the Benjamini-Hochberg procedure across the 12 comparisons (5% false discovery rate). Excluded results for *S. stercoralis* (NIE) because seroprevalence was low (<1%). WASH=combined water, sanitation and handwashing; Diff=difference; 95% CI=95% confidence interval; FDR=False Discovery Rate.

## Discussion

In this longitudinal analysis of the WASH Benefits Bangladesh trial, IgG seroprevalence increased sharply by age 2 years for many enteric pathogens, consistent with high transmission and accumulating pathogen exposure during early childhood. This study helps fill an important gap in WASH trials: stool-based PCR provides evidence of active infection at time of sampling, but infrequent stool collection can miss intermittent, short-duration or earlier infections. By measuring IgG responses to 12 enteric pathogens, we were able to characterise cumulative pathogen-specific exposure and estimate how WASH and nutrition interventions affected force of infection over the first two years of life. Over 94% of children 24 months old were seropositive for the highest transmission enteropathogens, including *Norovirus* GI/GII, LT-ETEC, and *Campylobacter*, with no benefit provided by WASH or nutrition interventions. Improved WASH led to modest reductions in seroprevalence to *Shigella/*EIEC, non-typhoidal *S. enterica*, *Cryptosporidium* and *Giardia*. These results are consistent with an overall reduction in enteric pathogen transmission and caregiver-reported symptomatic diarrhoea among children born into WASH compounds, even though a majority of children in intervention compounds were IgG seropositive to ≥5 pathogens before their second birthday. Improvements in household WASH conditions likely reduced enteric pathogen transmission broadly, and may have delayed the timing of primary infections, but did not entirely prevent enteric pathogen infection during the study. The PAASIM and MapSan Mozambique controlled trials found no overall neighbourhood and compound-level WASH effect on enteric pathogen infections at 12 and 24 after intervention, respectively, in an urban setting.^22,23^ However, exploratory analysis provided suggestive evidence of reduced *Trichuris* and *Shigella* infections among children born into the study by 24-month follow up visit suggesting that WASH may have delayed pathogen exposure and accumulation of enteric infections.^22^

These findings add to the broader body of evidence from the WASH Benefits Bangladesh trial by showing how serology can complement single-timepoint stool-based assessments of enteric pathogen infection. Earlier analyses using PCR^9,11^ and microscopy^10^ found mixed effects of WASH across pathogens, with limited reductions in most bacterial and some parasitic infections but evidence of reductions in selected enteric viruses, including *norovirus*. This IgG analysis suggests modest WASH-associated reductions in cumulative exposure to selected bacterial and protozoan pathogens, including *Shigella*/EIEC, *S. enterica*, *Cryptosporidium*, and *Giardia*. Together, these findings demonstrate that WASH can interrupt multiple faecal-oral transmission pathways and may reduce transmission across broad pathogen taxa. However, the magnitude of these differences was generally modest, consistent with household-level interventions being protective but insufficient to substantially disrupt enteric pathogen transmission in this setting. The timing of infection also appears important: WASH effects on enteric pathogen infection outcomes were more apparent during the second year of life, when children likely have greater mobility, environmental contact, and cumulative exposure risk. However, this pattern should be interpreted cautiously because the visit 2 analysis was limited to the EED sub-study and had lower statistical power.

The reduction in *Shigella*/EIEC IpaB seroprevalence associated with improved WASH contrasts with a recent analysis that found no effect of combined WASH + Nutrition on *Shigella*/EIEC IgG immune repertoire development measured through a bacterial display assay.^24^ Differences could arise from differences in analytic sample and assay. The prior analysis compared the combined WASH + Nutrition arm with the control arm in a subset of 120 children, whereas this analysis used a larger subset of the birth cohort (n=2453 samples) and compared children who received WASH with those who did not within the factorial design. Additionally, the prior analysis used peptide arrays to tile epitope-level IgG responses across several proteins (IpaA, IpaB, IpaC, IpaH) with relatively muted IpaB response and may have not been able to distinguish the modest reduction in transmission measured here on the Luminex platform. Further analyses using the bacterial display assay found poor alignment with the Luminex IgG responses for proteins with conformational epitopes with additional (potential) challenges of non-specific epitope binding, though overall age-dependent patterns for high transmission *Norovirus* GI/GII were broadly consistent between the assays.^25^

Our findings also highlight both the value and the limitations of serology for measuring enteric pathogen transmission in early childhood. IgG responses integrate exposure over a longer period and can provide a clearer picture of the cumulative force of infection through age-seroprevalence curves. This was particularly informative for pathogens with substantial levels of seroconversion that had not yet saturated near 100% by age 2 years, such as *Shigella*, *Cryptosporidium*, and *Giardia*. For example, prior PCR-based analyses found that *Cryptosporidium* infections were too rare at the final visit to support robust arm comparisons^9^, whereas IgG responses provided sufficient information to compare cumulative exposure across intervention groups. However, the same feature that makes IgG useful for capturing cumulative exposure can limit interpretation for pathogens with extremely high force of infection, where asymptomatic carriage of multiple enteric pathogens is common and children may be repeatedly exposed over time.^26,27^ Prior serologic studies, including work on *norovirus* in young children in low-resource settings, have similarly shown that cumulative exposure can be substantially higher ^2,28,29^ than suggested by PCR prevalence at a single visit. In our study, the high *Norovirus* GI/GII seroprevalence (∼80%) at 14 months contrasted with lower PCR point prevalence (∼23%)^11^, underscoring how serology can provide a broader picture of cumulative burden. Thus, PCR may be preferable for detecting short-term reductions in infection for very high-transmission pathogens, while serology may be especially useful for estimating cumulative burden and intervention effects on pathogens for which seroprevalence remains in a more intermediate range, below 100%.

The nutrition findings should be interpreted alongside prior trial results showing a ∼40% reduction in diarrhoea in the nutrition-alone arm.^8^ Although the nutrition package included lipid-based nutrient supplements and breastfeeding promotion, which could plausibly reduce exposure or susceptibility to enteric infection, we found no broad reductions in pathogen-specific seroprevalence. Breastfeeding was already highly prevalent across groups at follow-up, leaving limited scope for the intervention to further increase breastfeeding practices. The most pronounced reduction was for non-typhoidal *S. enterica* seroprevalence at the 14-month visit, near the end of the breastfeeding promotion, while effects for other pathogens were small and not statistically significant. Together with no evidence of reduction in PCR-detected enteric pathogens due to nutrition alone^9,11^ these findings suggest that the large reduction in diarrhoea in the nutrition group may reflect courtesy bias.

Our findings support previous evidence that children remained highly exposed to faecal contamination and enteric pathogens^30^ despite good adherence to household WASH interventions^31^ and some reduction in diarrhoea or enteric infections.^32^ These findings raise questions about whether basic household WASH interventions can adequately interrupt diverse transmission pathways, including foodborne, animal faecal, person-to-person and broader environmental exposure routes.^33^ The results also underscore the importance of directly measuring enteric pathogen infection, rather than relying only on behavioural, process or environmental indicators or self-reported health outcomes.

This study has several limitations. Serologic measures provide integrated markers of cumulative pathogen exposure, but they do not indicate active infection, pathogen shedding, or the precise timing of infection because IgG responses may persist after exposure. Seroprevalence may also reflect variation in host antibody responses, not exposure alone. In particular, nutrition, breastfeeding, and immune development potentially related to the early-life microbiome may alter systemic antibody responses following exposure; however, microbiome composition was not measured in this study. Serologic saturation for highly prevalent pathogens also limited our ability to evaluate intervention effects for pathogens with very high transmission intensity. DBS measurements may be affected by variation in specimen quality, storage conditions, or antibody recovery, although laboratory quality-control procedures were used to reduce potential measurement error. Since DBS specimens were unavailable for some eligible children, the complete case analysis assumed outcomes missing completely at random. We also assessed IgG responses, and some mucosal pathogens are non-invasive (such as *V. cholerae*) and largely induce IgA and IgM responses.^34^ Induction of systemic IgG responses in such cases often requires repetitive exposures, and as such our results may underrepresent force of infection compared to those relating to invasive enteropathy (example *S. enterica*, *Campylobacter*, *Shigella*/EIEC). In addition, the trial was conducted in low-resource rural communities in Bangladesh, and cluster selection based on logistical feasibility may limit generalizability to settings with different transmission intensity, intervention coverage, or environmental conditions.

This study also has important strengths. We leveraged a rigorously conducted cluster-randomized trial with high intervention adherence and longitudinal follow-up during a critical period of early childhood. Multiplex IgG testing provided an efficient way to assess overall pathogen transmission and pathogen-specific impacts of the interventions in a trial with infrequent specimen collection. By complementing prior PCR and microscopy analyses from the same trial, this study provides new insight into how WASH may affect cumulative pathogen exposure over time.

In conclusion, longitudinal multiplex serology provided new evidence on early-life exposure to multiple enteric pathogens in the WASH Benefits Bangladesh trial. Although WASH effects were modest and pathogen-specific, the generally protective pattern for multiple bacterial and protozoan pathogens suggest that improved WASH may reduce cumulative exposure to some enteric pathogens over time, but insufficient to substantially disrupt transmission in this high-exposure setting. These findings support the use of serology as a complementary tool for evaluating enteric pathogen transmission and intervention effects in large-scale trials, particularly low-to-medium prevalence pathogens, while underscoring the need to combine serologic and stool-based measures to capture both cumulative exposure and short-term infection dynamics.

## Data Availability

The pre-analysis plan, de-identified data, and analysis scripts to reproduce all analyses will be available through Zenodo (https://doi.org/10.5281/zenodo.23671136).

## Contributors

Conceptualization: MR, PAA-T, PJL, RH, JWP, BFA, SMN. Data curation: RWK, HH, PAA-T, EK, SJB, ANM, BFA. Formal analysis: PAA-T, EK, HH, SJB, BFA. Funding acquisition: MR, RH, ETR, CPS, AJP, CN, JMC, SPL, PJL, JWP, BFA, SMN. Investigation: HK, RWK, HH, HE, SAli, MZR, AKS, MRM, SAshraf, SMP, PAA-T, BFA, SMN. Methodology: HH, PAA-T, EK, SJB, JBC, NW, JWP, BFA, SMN. Project administration: MR, SAli, MZR, AKS, MRM, SAshraf, SMP, JG, AL. Resources: SAli, MZR, AKS, MRM, SAshraf, SMP, HK, RWK, HE, EBG, CC, MMS, JG, RCC, ETR, CM, RLA, AL, JWP. Software: PAA-T, HH, EK, SJB, JC, BFA. Supervision: MR, HK, EBG, CC, CPS, JG, JBC, AE, AL, JMC, SPL, RH, BFA, SMN. Validation: RWK, HE, EBG, CC, RCC, CM, AKS. Visualization: HH, PAA-T, SJB, JC. Writing – original draft: MR, PAA-T, BFA. Writing – review & editing: All authors.

## Data sharing

The pre-analysis plan, de-identified data, and analysis scripts to reproduce all analyses will be available through Zenodo (https://doi.org/10.5281/zenodo.23671136). All analyses were conducted in R (version 4.3.3).

## Acknowledgments

We are sincerely grateful to Diana Martin and Gretchen Cooley (US CDC) for guidance with the multiplex bead assay, to Jan Vinje (US CDC) for assistance with norovirus targets, and to William Petri (University of Virginia) and Joel Herbein (TechLab) for donating LecA. icddr,b is also grateful to the Govt. of Bangladesh and Canada for providing core/unrestricted support.

## Use of generative AI and AI-assisted technologies

Claude Opus 5.5 (Anthropic) assisted with Supplementary Table 2, R code review, and data/code documentation. The authors reviewed all outputs and take full responsibility.

