## Supplementary Materials for "Effects of water quality, sanitation, handwashing and nutritional interventions on enteric pathogen seroconversion in rural Bangladesh: a cluster randomised trial"

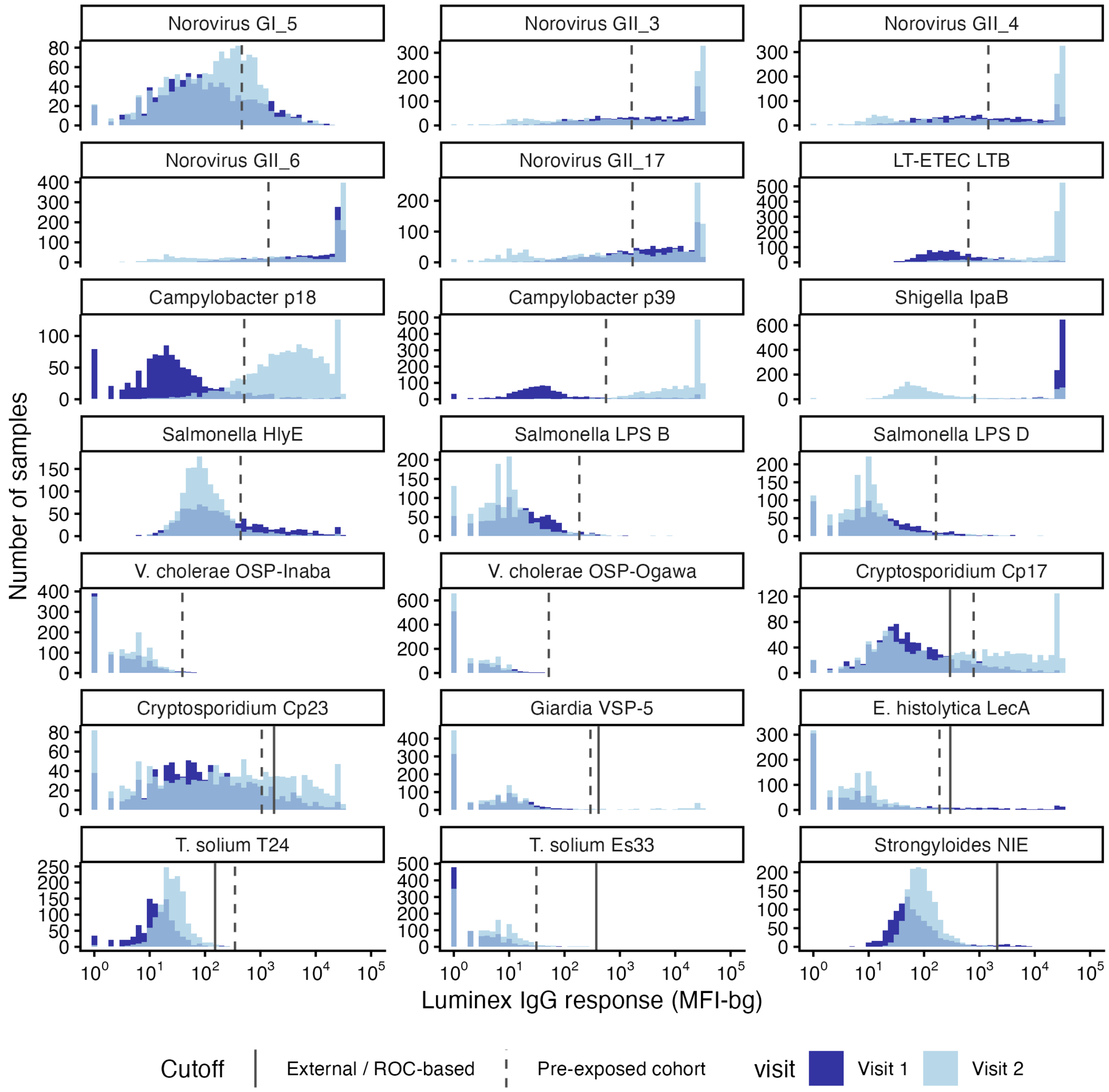


**Supplementary Figure 1. Luminex IgG response (MFI-bg) to enteric pathogens during visits 1 and 2 in the WASH Benefits Bangladesh environmental enteric dysfunction substudy.** Children were median ages 3 months (visit 1) and 14 months (visit 2) at the time of measurement. Vertical lines mark seropositivity cutoffs derived using external samples (solid) for protozoan and helminth pathogens, and from longitudinal boosting in a pre-exposed cohort approach (dashed).

**Supplementary Table 1: Baseline characteristics by intervention groups.**

| | Control (N=897) | WASH (N=534) | Nutrition (N=495) | WASH + Nutrition (N=527) |
|---|---|---|---|---|
| **Maternal age (years)** Median [Min, Max] | 23 [15, 43] | 25 [15, 44] | 23 [15, 45] | 23 [14, 43] |
| **Maternal education** No education | 119 (13.3%) | 70 (13.1%) | 81 (16.5%) | 82 (15.6%) |
| Primary (1-5y) | 280 (31.4%) | 172 (32.2%) | 145 (29.5%) | 167 (31.7%) |
| Secondary (>5y) | 493 (55.3%) | 292 (54.7%) | 266 (54.1%) | 278 (52.8%) |
| **Maternal height (cm)** Median [Min, Max] | 151 [134, 167] | 150 [136, 164] | 150 [127, 168] | 150 [133, 167] |
| **Paternal education** No education | 252 (28.3%) | 164 (30.9%) | 150 (30.5%) | 157 (29.8%) |
| Primary (1-5y) | 265 (29.8%) | 152 (28.6%) | 155 (31.6%) | 170 (32.3%) |
| Secondary (>5y) | 373 (41.9%) | 215 (40.5%) | 186 (37.9%) | 199 (37.8%) |
| **Works in agriculture** | 273 (30.6%) | 160 (30.0%) | 167 (33.9%) | 162 (30.8%) |
| **International wealth index** Median [Min, Max] | 33.7 [0.00, 63.0] | 31.7 [4.66, 63.0] | 29.6 [0, 63.0] | 31.5 [4.66, 63.0] |
| **Number of households per compound** Mean (SD) | 11.0 (6.17) | 11.1 (5.90) | 11.6 (6.51) | 11.0 (6.48) |
| **Number of people per compound** Mean (SD) | 4.73 (2.23) | 4.62 (2.06) | 4.65 (2.10) | 4.76 (2.12) |
| **Primary drinking water source (tubewell)** | 691 (77.5%) | 424 (79.4%) | 367 (74.6%) | 391 (74.2%) |
| **Distance to primary water source (min)** Mean (SD) | 1.29 (20.1) | 0.979 (5.37) | 0.616 (1.29) | 0.706 (1.99) |
| **Reported storing drinking water** | 404 (45.3%) | 222 (41.6%) | 209 (42.5%) | 253 (48.0%) |
| **Reported treating water today or yesterday** | 3 (0.3%) | 0 (0%) | 0 (0%) | 2 (0.4%) |
| **Latrine** No latrine | 408 (48.6%) | 245 (48.1%) | 219 (47.1%) | 245 (49.6%) |
| Latrine without water seal | 267 (31.8%) | 184 (36.1%) | 161 (34.6%) | 174 (35.2%) |
| Latrine with water seal | 164 (19.5%) | 80 (15.7%) | 85 (18.3%) | 75 (15.2%) |
| **Primary handwashing location has water and soap** | 190 (22.9%) | 119 (24.2%) | 99 (21.9%) | 121 (24.6%) |
| **HFIAS food insecurity** Moderate/severe | 201 (22.5%) | 118 (22.1%) | 104 (21.1%) | 105 (19.9%) |

WASH=combined water, sanitation and handwashing; SD=standard deviation; HFIAS=Household Food Insecurity Access Scale.

**Supplementary Table 2: Measurement of intervention adherence by study group at enrollment, year 1 and year 2 follow-up.**

| Indicator | Round | WASH | No WASH | Nutrition | No Nutrition |
|---|---|---|---|---|---|
| Number of compounds assessed | Enrollment | 1388/1388 (100%) | 2081/2081 (100%) | 1385/1385 (100%) | 2084/2084 (100%) |
| | Year 1 | 1205/1388 (87%) | 1732/2081 (83%) | 1181/1385 (85%) | 1756/2084 (84%) |
| | Year 2 | 1174/1388 (85%) | 1712/2081 (82%) | 1160/1385 (84%) | 1726/2084 (83%) |
| Stored drinking water | Enrollment | 635/1388 (46%) | 967/2081 (46%) | 632/1385 (46%) | 970/2084 (47%) |
| | Year 1 | 1165/1205 (97%) | 732/1732 (42%) | 806/1181 (68%) | 1091/1756 (62%) |
| | Year 2 | 1127/1174 (96%) | 710/1712 (41%) | 794/1160 (68%) | 1043/1726 (60%) |
| Stored drinking water has detectable free chlorine (>0.1 mg/L) | Enrollment | .. | .. | .. | .. |
| | Year 1 | 939/1180 (80%) | .. | .. | .. |
| | Year 2 | 972/1157 (84%) | .. | .. | .. |
| Latrine with a functional water seal | Enrollment | 307/1155 (27%) | 541/1758 (31%) | 338/1160 (29%) | 510/1753 (29%) |
| | Year 1 | 1137/1202 (95%) | 457/1579 (29%) | 713/1130 (63%) | 881/1651 (53%) |
| | Year 2 | 1128/1170 (96%) | 487/1579 (31%) | 724/1109 (65%) | 891/1640 (54%) |
| No visible feces on latrine slab or floor | Enrollment | 587/1296 (45%) | 956/1956 (49%) | 629/1292 (49%) | 914/1960 (47%) |
| | Year 1 | 1049/1203 (87%) | 991/1643 (60%) | 860/1153 (75%) | 1180/1693 (70%) |
| | Year 2 | 979/1170 (84%) | 925/1626 (57%) | 808/1124 (72%) | 1096/1672 (66%) |
| Handwashing location has soap | Enrollment | 304/1286 (24%) | 446/1900 (23%) | 301/1284 (23%) | 449/1902 (24%) |
| | Year 1 | 1082/1204 (90%) | 455/1515 (30%) | 708/1111 (64%) | 829/1608 (52%) |
| | Year 2 | 1071/1174 (91%) | 515/1712 (30%) | 735/1160 (63%) | 851/1726 (49%) |
| Lipid Nutrient Supplement sachets consumed (% expected)* | Enrollment | .. | .. | .. | .. |
| | Year 1 | 94% | 93% | 94% | .. |
| | Year 2 | 93% | 94% | 93% | .. |

Data are n (%) or %.
'..' = not measured or not interpretable on this margin.
Free chlorine reported for the WASH group only; *LNS % are conditional on being in a Nutrition cluster. Both not measured at enrolment.

**Supplemental Table 3. Enteric pathogen seroprevalence at the 24-month visit by individual treatment group and main-effect comparison.**

| Pathogen | Intervention group | Seroprevalence n/N (%) | Main effect group | Seroprevalence n/N (%) | RD (95% CI) | p-value |
|---|---|---|---|---|---|---|
| **Viruses** | | | | | | |
| ***Norovirus* GI/GII** | Control | 863/897 (96.2%) | No WASH | 1336/1391 (96%) | Ref | — |
| | WASH | 515/534 (96.4%) | WASH | 1029/1061 (97%) | 0.9 (-0.3, 2.1) | 0.229 |
| | Nutrition | 473/494 (95.7%) | No Nutrition | 1378/1431 (96.3%) | Ref | — |
| | WASH + Nutrition | 514/527 (97.5%) | Nutrition | 987/1021 (96.7%) | 0.3 (-0.9, 1.5) | 0.700 |
| **Bacteria** | | | | | | |
| **LT-ETEC (LTB)** | Control | 891/896 (99.4%) | No WASH | 1382/1391 (99.4%) | Ref | — |
| | WASH | 531/534 (99.4%) | WASH | 1053/1061 (99.2%) | -0.0 (-0.6, 0.5) | 0.936 |
| | Nutrition | 491/495 (99.2%) | No Nutrition | 1422/1430 (99.4%) | Ref | — |
| | WASH + Nutrition | 522/527 (99.1%) | Nutrition | 1013/1022 (99.1%) | -0.3 (-0.8, 0.3) | 0.375 |
| ***Campylobacter* (p18,p39)** | Control | 827/896 (92.3%) | No WASH | 1292/1391 (92.9%) | Ref | — |
| | WASH | 499/534 (93.4%) | WASH | 996/1061 (93.9%) | 0.8 (-1.0, 2.6) | 0.467 |
| | Nutrition | 465/495 (93.9%) | No Nutrition | 1326/1430 (92.7%) | Ref | — |
| | WASH + Nutrition | 497/527 (94.3%) | Nutrition | 962/1022 (94.1%) | 1.2 (-0.6, 3.0) | 0.287 |
| ***Shigella*/EIEC (IpaB)** | Control | 719/896 (80.2%) | No WASH | 1099/1391 (79%) | Ref | — |
| | WASH | 397/534 (74.3%) | WASH | 790/1061 (74.5%) | -4.5 (-7.6, -1.5) | 0.011 |
| | Nutrition | 380/495 (76.8%) | No Nutrition | 1116/1430 (78%) | Ref | — |
| | WASH + Nutrition | 393/527 (74.6%) | Nutrition | 773/1022 (75.6%) | -1.5 (-4.5, 1.6) | 0.402 |
| ***S. enterica* typhi (HlyE)** | Control | 107/896 (11.9%) | No WASH | 176/1391 (12.7%) | Ref | — |
| | WASH | 83/534 (15.5%) | WASH | 156/1061 (14.7%) | 2.1 (-0.5, 4.6) | 0.177 |
| | Nutrition | 69/495 (13.9%) | No Nutrition | 190/1430 (13.3%) | Ref | — |
| | WASH + Nutrition | 73/527 (13.9%) | Nutrition | 142/1022 (13.9%) | 0.2 (-2.4, 2.9) | 0.888 |
| ***S. enterica* (LPS B,D)** | Control | 136/896 (15.2%) | No WASH | 211/1390 (15.2%) | Ref | — |
| | WASH | 66/534 (12.4%) | WASH | 118/1061 (11.1%) | -4.0 (-6.5, -1.5) | 0.008 |
| | Nutrition | 75/494 (15.2%) | No Nutrition | 202/1430 (14.1%) | Ref | — |
| | WASH + Nutrition | 52/527 (9.9%) | Nutrition | 127/1021 (12.4%) | -1.3 (-3.9, 1.3) | 0.389 |
| ***V. cholerae* (OSP-Inaba/Ogawa)** | Control | 29/896 (3.2%) | No WASH | 46/1390 (3.3%) | Ref | — |
| | WASH | 15/534 (2.8%) | WASH | 27/1061 (2.5%) | -0.8 (-2.2, 0.5) | 0.292 |
| | Nutrition | 17/494 (3.4%) | No Nutrition | 44/1430 (3.1%) | Ref | — |
| | WASH + Nutrition | 12/527 (2.3%) | Nutrition | 29/1021 (2.8%) | -0.2 (-1.6, 1.1) | 0.752 |

| | | | | | | |
|---|---|---|---|---|---|---|
| **Protozoa** | | | | | | |
| ***Cryptosporidium* (Cp17,Cp23)** | Control | 562/896 (62.7%) | No WASH | 892/1391 (64.1%) | Ref | — |
| | WASH | 319/534 (59.7%) | WASH | 660/1061 (62.2%) | -2.9 (-6.5, 0.7) | 0.173 |
| | Nutrition | 330/495 (66.7%) | No Nutrition | 881/1430 (61.6%) | Ref | — |
| | WASH + Nutrition | 341/527 (64.7%) | Nutrition | 671/1022 (65.7%) | 4.1 (0.4, 7.8) | 0.051 |
| ***Giardia* (VSP-5)** | Control | 286/897 (31.9%) | No WASH | 443/1391 (31.8%) | Ref | — |
| | WASH | 153/534 (28.7%) | WASH | 300/1061 (28.3%) | -3.4 (-6.7, -0.2) | 0.069 |
| | Nutrition | 157/494 (31.8%) | No Nutrition | 439/1431 (30.7%) | Ref | — |
| | WASH + Nutrition | 147/527 (27.9%) | Nutrition | 304/1021 (29.8%) | -0.4 (-3.7, 2.8) | 0.820 |
| ***E. histolytica* (LecA)** | Control | 13/896 (1.5%) | No WASH | 24/1391 (1.7%) | Ref | — |
| | WASH | 7/534 (1.3%) | WASH | 22/1061 (2.1%) | 0.2 (-0.7, 1.2) | 0.703 |
| | Nutrition | 11/495 (2.2%) | No Nutrition | 20/1430 (1.4%) | Ref | — |
| | WASH + Nutrition | 15/527 (2.8%) | Nutrition | 26/1022 (2.5%) | 1.0 (0.1, 2.0) | 0.052 |
| **Helminths** | | | | | | |
| ***T. solium* (T24,Es33)** | Control | 19/896 (2.1%) | No WASH | 36/1390 (2.6%) | Ref | — |
| | WASH | 15/534 (2.8%) | WASH | 32/1061 (3%) | 0.2 (-1.0, 1.4) | 0.739 |
| | Nutrition | 17/494 (3.4%) | No Nutrition | 34/1430 (2.4%) | Ref | — |
| | WASH + Nutrition | 17/527 (3.2%) | Nutrition | 34/1021 (3.3%) | 0.9 (-0.3, 2.1) | 0.195 |
| ***S. stercoralis* (NIE)** | Control | 1/896 (0.1%) | No WASH | 2/1391 (0.1%) | Ref | — |
| | WASH | 1/534 (0.2%) | WASH | 3/1061 (0.3%) | 0.1 (-0.2, 0.4) | 0.616 |
| | Nutrition | 1/495 (0.2%) | No Nutrition | 2/1430 (0.1%) | Ref | — |
| | WASH + Nutrition | 2/527 (0.4%) | Nutrition | 3/1022 (0.3%) | 0.1 (-0.2, 0.4) | 0.633 |

Note:
n/N (%) shows observed seropositive counts and percentages.
Risk differences are marginal main-effect estimates reported in percentage points.
P values are two-sided permutation p-values from the primary main-effect models.

| Pathogen | Nutrition | No Nutrition | Diff (95% CI) | p-value | FDR p-value |
|---|---|---|---|---|---|
| Norovirus GI/GII | 96.7% | 96.4% | 0.3 (-0.9, 1.5) | 0.70 | 0.89 |
| LT-ETEC (LTB) | 99.2% | 99.4% | -0.3 (-0.8, 0.3) | 0.38 | 0.69 |
| Campylobacter (p18,p39) | 94.0% | 92.8% | 1.2 (-0.6, 3.0) | 0.29 | 0.69 |
| Shigella/EIEC (IpaB) | 75.9% | 77.4% | -1.5 (-4.5, 1.6) | 0.40 | 0.69 |
| S. enterica typhi (HlyE) | 14.1% | 13.9% | 0.2 (-2.4, 2.9) | 0.89 | 0.89 |
| S. enterica (LPS B,D) | 12.5% | 13.9% | -1.3 (-3.9, 1.3) | 0.39 | 0.69 |
| V. cholerae (OSP-Inaba/Ogawa) | 2.8% | 3.1% | -0.2 (-1.6, 1.1) | 0.75 | 0.89 |
| Cryptosporidium (Cp17,Cp23) | 65.4% | 61.2% | 4.1 (0.4, 7.8) | 0.05 | 0.31 |
| Giardia (VSP-5) | 30.0% | 30.5% | -0.4 (-3.7, 2.8) | 0.82 | 0.89 |
| E. histolytica (LecA) | 2.4% | 1.4% | 1.0 (0.1, 2.0) | 0.05 | 0.31 |
| T. solium (T24,Es33) | 3.3% | 2.4% | 0.9 (-0.3, 2.1) | 0.20 | 0.69 |

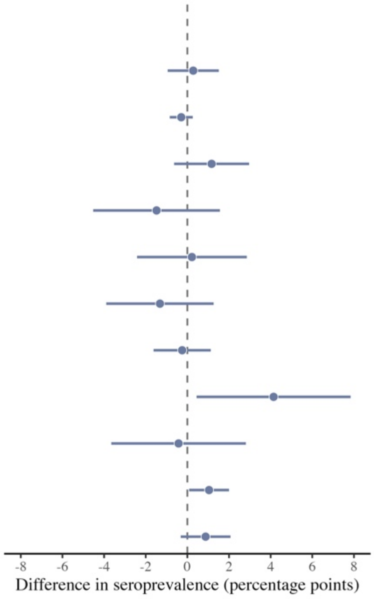


**Supplementary Figure 2. Nutrition intervention effects on enteric pathogen seroprevalence at median age 24 months.** Effects estimated as marginal means and treatment effects for clusters that received Nutrition (Nutrition, WASH + Nutrition) compared with clusters that did not (WASH, Control)**.** Two-sided permutation p-values were estimated by permuting treatment assignments (10,006 replicates). P-values were adjusted for multiple comparisons using the Benjamini-Hochberg procedure across the 12 comparisons (5% false discovery rate). Excluded results for *S. stercoralis (NIE)* because seroprevalence was low (<1%).

WASH=combined water, sanitation and handwashing; Diff=difference; 95% CI=95% confidence interval; FDR=False Discovery Rate.

**A**

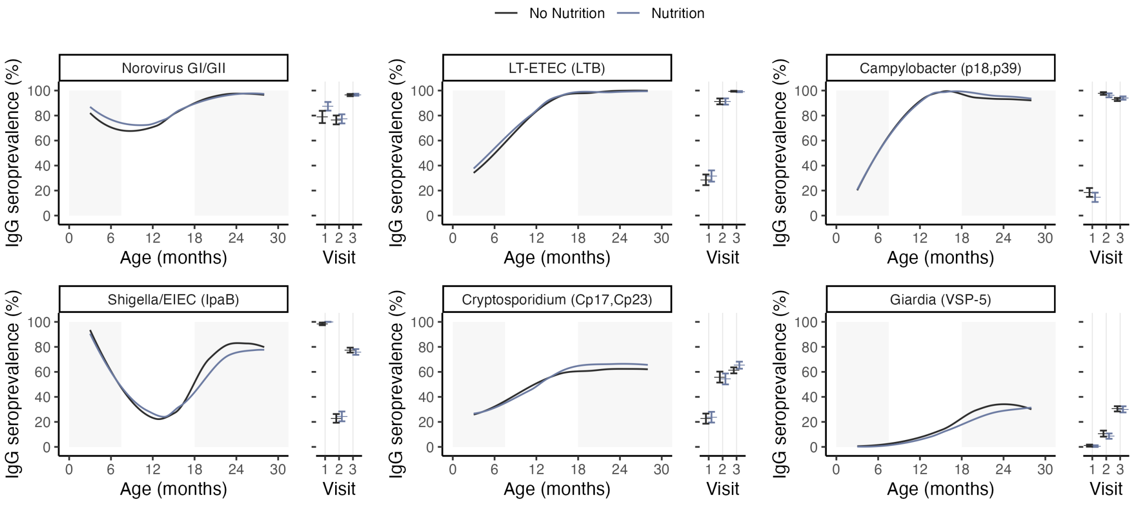


**B**

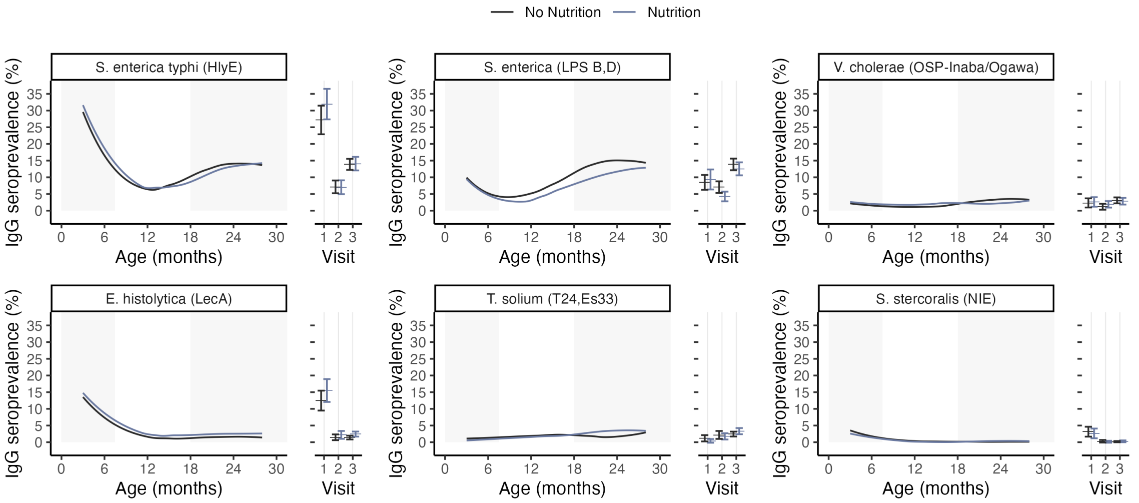


**Supplementary Figure 3. Enteric pathogen seroprevalence by age and Nutrition intervention group, Bangladesh 2013-2016. A.** Higher transmission pathogens. **B.** lower transmission pathogens. The difference between panels is the Y-axis range, to better display the data. Lines represent age-seroprevalence estimated by treatment group using locally weighted regression over individual seropositivity data (points), trimmed to ages 3-28 months to avoid edge effects. Points and error bars to the right of each panel show treatment-specific marginal average seroprevalence and 95% confidence intervals by study visit (1, 2, 3). Gray shading shows child age ranges of visits 1 and 3, with the primary outcome and full cohort measured at visit 3. Similar estimates summarized by the WASH intervention margin are in **Figure 3**. WASH=combined water, sanitation and handwashing intervention.

**Supplementary Table 4. Enteric pathogen seroprevalence at the 14-month visit by individual treatment group and main-effect comparison**

| Pathogen | Intervention group | Seroprevalence n/N (%) | Main effect group | Seroprevalence n/N (%) | RD (95% CI) | p-value |
|---|---|---|---|---|---|---|
| **Viruses** | | | | | | |
| ***Norovirus* GI/GII** | Control | 77.9 (250/321) | No WASH | 78.9 (520/659) | Ref | — |
| | WASH | 75.1 (274/365) | WASH | 75 (533/711) | -3.9 (-9.0, 1.2) | 0.59 |
| | Nutrition | 79.9 (270/338) | No Nutrition | 76.4 (524/686) | Ref | — |
| | WASH + Nutrition | 74.9 (259/346) | Nutrition | 77.3 (529/684) | 0.9 (-4.2, 5.9) | 0.89 |
| **Bacteria** | | | | | | |
| **LT-ETEC (LTB)** | Control | 93.5 (300/321) | No WASH | 92.3 (608/659) | Ref | — |
| | WASH | 89.3 (326/365) | WASH | 90.3 (642/711) | -2.0 (-5.4, 1.4) | 0.62 |
| | Nutrition | 91.1 (308/338) | No Nutrition | 91.3 (626/686) | Ref | — |
| | WASH + Nutrition | 91.3 (316/346) | Nutrition | 91.2 (624/684) | -0.1 (-3.5, 3.3) | 0.97 |
| ***Campylobacter* (p18,p39)** | Control | 97.5 (313/321) | No WASH | 96.8 (638/659) | Ref | — |
| | WASH | 97.8 (357/365) | WASH | 97 (690/711) | 0.2 (-1.7, 2.1) | 0.97 |
| | Nutrition | 96.2 (325/338) | No Nutrition | 97.7 (670/686) | Ref | — |
| | WASH + Nutrition | 96.2 (333/346) | Nutrition | 96.2 (658/684) | -1.5 (-3.4, 0.5) | 0.69 |
| ***Shigella*/EIEC (IpaB)** | Control | 21.5 (69/321) | No WASH | 22.6 (149/659) | Ref | — |
| | WASH | 24.1 (88/365) | WASH | 24.6 (175/711) | 2.0 (-3.2, 7.3) | 0.66 |
| | Nutrition | 23.7 (80/338) | No Nutrition | 22.9 (157/686) | Ref | — |
| | WASH + Nutrition | 25.1 (87/346) | Nutrition | 24.4 (167/684) | 1.6 (-3.7, 6.8) | 0.86 |
| ***S. enterica typhi* (HlyE)** | Control | 7.5 (24/321) | No WASH | 7 (46/659) | Ref | — |
| | WASH | 6.8 (25/365) | WASH | 7.2 (51/711) | 0.2 (-2.6, 3.0) | 0.97 |
| | Nutrition | 6.5 (22/338) | No Nutrition | 7.1 (49/686) | Ref | — |
| | WASH + Nutrition | 7.5 (26/346) | Nutrition | 7 (48/684) | -0.1 (-2.9, 2.7) | 0.97 |
| ***S. enterica* (LPS B,D)*** | Control | 9.7 (31/321) | No WASH | 6.4 (42/659) | Ref | — |
| | WASH | 4.7 (17/365) | WASH | 4.9 (35/711) | -5.0 (8.4, -1.6) | 0.05 |
| | Nutrition | 3.3 (11/338) | No Nutrition | 7 (48/686) | Ref | — |
| | WASH + Nutrition | 5.2 (18/346) | Nutrition | 4.2 (29/684) | -6.4 (-9.8, -3.0) | 0.00 |
| ***V. cholerae* (OSP-Inaba/Ogawa)** | Control | 1.9 (6/321) | No WASH | 1.8 (12/659) | Ref | — |
| | WASH | 0.5 (2/365) | WASH | 1.3 (9/711) | -0.5 (-1.9, 0.8) | 0.66 |
| | Nutrition | 1.8 (6/338) | No Nutrition | 1.2 (8/686) | Ref | — |
| | WASH + Nutrition | 2 (7/346) | Nutrition | 1.9 (13/684) | 0.7 (-0.6, 2.1) | 0.69 |

| | | | | | | |
|---|---|---|---|---|---|---|
| **Protozoa** | | | | | | |
| ***Cryptosporidium* (Cp17,Cp23)** | Control | 53 (170/321) | No WASH | 53.1 (350/659) | Ref | — |
| | WASH | 58.6 (214/365) | WASH | 57.1 (406/711) | 4.0 (-2.2, 10.1) | 0.62 |
| | Nutrition | 53.3 (180/338) | No Nutrition | 56 (384/686) | Ref | — |
| | WASH + Nutrition | 55.5 (192/346) | Nutrition | 54.4 (372/684) | -1.5 (-3.4, 0.5) | 0.86 |
| ***Giardia* (VSP-5)** | Control | 10.3 (33/321) | No WASH | 9.3 (61/659) | Ref | — |
| | WASH | 11 (40/365) | WASH | 10 (71/711) | 0.7 (-2.5, 3.9) | 0.91 |
| | Nutrition | 8.3 (28/338) | No Nutrition | 10.6 (73/686) | Ref | — |
| | WASH + Nutrition | 9 (31/346) | Nutrition | 8.6 (59/684) | -2.0 (-5.2, 1.2) | 0.69 |
| ***E. histolytica* (LecA)** | Control | 1.6 (5/321) | No WASH | 2.1 (14/659) | Ref | — |
| | WASH | 1.4 (5/365) | WASH | 1.5 (11/711) | -0.6 (-2.0, 0.9) | 0.66 |
| | Nutrition | 2.7 (9/338) | No Nutrition | 1.5 (10/686) | Ref | — |
| | WASH + Nutrition | 1.7 (6/346) | Nutrition | 2.2 (15/684) | 0.7 (-0.7, 2.2) | 0.69 |
| **Helminths** | | | | | | |
| ***T. solium* (T24,Es33)** | Control | 2.5 (8/321) | No WASH | 2 (13/659) | Ref | — |
| | WASH | 1.9 (7/365) | WASH | 2 (14/711) | 0.0 (-1.5, 1.5) | 0.99 |
| | Nutrition | 1.5 (5/338) | No Nutrition | 2.2 (15/686) | Ref | — |
| | WASH + Nutrition | 2 (7/346) | Nutrition | 1.8 (12/684) | -0.4 (-1.9, 1.1) | 0.86 |
| ***S. stercoralis* (NIE)*** | Control | 0 (0/321) | No WASH | 0.2 (1/659) | Ref | — |
| | WASH | 0.5 (2/365) | WASH | 0.3 (2/711) | 0.1 (-0.3, 0.6) | 0.99 |
| | Nutrition | 0.3 (1/338) | No Nutrition | 0.3 (2/686) | Ref | — |
| | WASH + Nutrition | 0 (0/346) | Nutrition | 0.1 (1/684) | 0.3 (-0.3, 0.9) | 0.69 |

Note:
n/N (%) shows observed seropositive counts and percentages.
Risk differences are marginal main-effect estimates reported in percentage points.
*Conditional means and effects are reported for pathogens with evidence of additive interaction.
P values are two-sided permutation p-values from the primary main-effect models.

| Pathogen | Nutrition | No Nutrition | Diff (95% CI) | p-value | FDR p-value |
|---|---|---|---|---|---|
| Norovirus GI/GII | 77.4% | 76.5% | 0.9 (-4.2, 5.9) | 0.74 | 0.89 |
| LT-ETEC (LTB) | 91.2% | 91.3% | -0.1 (-3.5, 3.3) | 0.97 | 0.97 |
| Campylobacter (p18,p39) | 96.2% | 97.7% | -1.5 (-3.4, 0.5) | 0.14 | 0.69 |
| Shigella/EIEC (IpaB) | 24.4% | 22.8% | 1.6 (-3.7, 6.8) | 0.56 | 0.86 |
| S. enterica typhi (HlyE) | 7.0% | 7.1% | -0.1 (-2.9, 2.7) | 0.93 | 0.97 |
| S. enterica (LPS B,D) * | 3.3% | 9.7% | -6.4 (-9.8, -3.0) | <0.01 | <0.01 |
| V. cholerae (OSP-Inaba/Ogawa) | 1.9% | 1.2% | 0.7 (-0.6, 2.1) | 0.29 | 0.69 |
| Cryptosporidium (Cp17,Cp23) | 54.4% | 55.8% | -1.5 (-7.7, 4.7) | 0.64 | 0.86 |
| Giardia (VSP-5) | 8.6% | 10.6% | -2.0 (-5.2, 1.2) | 0.23 | 0.69 |
| E. histolytica (LecA) | 2.2% | 1.5% | 0.7 (-0.7, 2.2) | 0.34 | 0.69 |
| T. solium (T24,Es33) | 1.8% | 2.2% | -0.4 (-1.9, 1.1) | 0.57 | 0.86 |

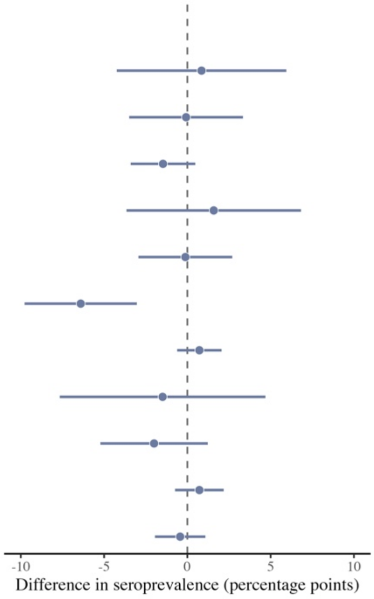


**Supplementary Figure 4. Nutrition intervention effects on enteric pathogen seroprevalence at median age 14 months.** Effects estimated as marginal means and treatment effects for clusters that received Nutrition (Nutrition, WASH + Nutrition) compared with clusters that did not (WASH, Control)**.** * Conditional means and effects comparing Nutrition vs Control are reported in cases of interaction between WASH and Nutrition interventions (P< 0.1) for *S. enterica* LPS Groups B and D. Two-sided permutation p-values were estimated by permuting treatment assignments (10,006 replicates). P-values were adjusted for multiple comparisons using the Benjamini-Hochberg procedure across the 12 comparisons (5% false discovery rate). Excluded results for *S. stercoralis* (NIE) because seroprevalence was low (<1%).
WASH=combined water, sanitation and handwashing; Diff=difference; 95% CI=95% confidence interval; FDR=False Discovery Rate.

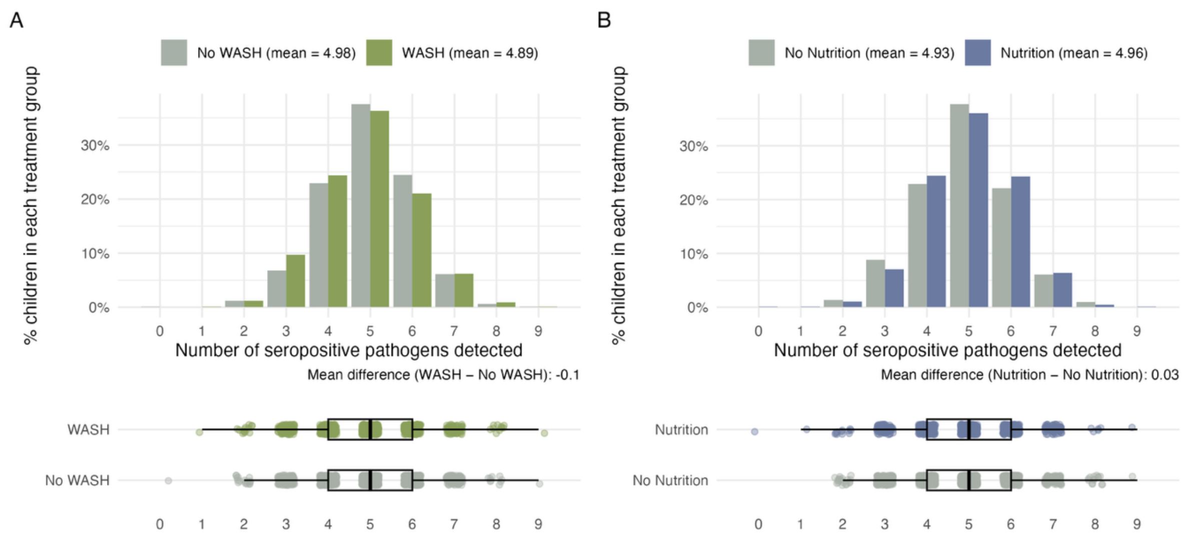


**Supplementary Figure 5. Distribution of the number of seropositive enteric pathogens detected among children by WASH and Nutrition interventions at median age 24 months.** WASH=combined water, sanitation and handwashing.
